# A composite biomarker for esophageal cancer risk from automated analysis of a non-endoscopic device

**DOI:** 10.1101/2021.08.20.21262366

**Authors:** Adam G. Berman, Rebecca C. Fitzgerald, Florian Markowetz

## Abstract

Barrett’s esophagus containing intestinal metaplasia predisposes to cancer, yet the majority of cases are undiagnosed. The length of a Barrett’s segment is a key indicator of cancer risk, but measuring it has so far relied on endoscopy, which is expensive and invasive. Cytosponge-TFF3 is a minimally-invasive test that identifies intestinal metaplasia for endoscopic confirmation. We report a machine learning technique to quantify the extent of intestinal metaplasia and predict Barrett’s segment length from whole-slide image tile counts automatically generated from Cytosponge-TFF3 histology slides. Utilizing data from 529 patients, our segment length prediction model achieves an average validation fold accuracy of 0.84. Applying this algorithm to an independent test set of 162 patients from a screening trial shows a precision of 0.90 for identifying short-segment disease. This advance will enable higher-risk patients to be prioritized for endoscopy while saving more than half of Cytosponge-TFF3-positive patients from endoscopy in the screening setting.

## Main

Esophageal adenocarcinoma (EAC) is the sixth leading cause of cancer-related death with an abysmal 5-year survival of 13%, usually only manifesting at a late stage (*1, 2*). Barrett’s esophagus (BE) is a precursor to EAC and its high prevalence in the Western world (*3, 4*) makes it an attractive target for early detection of EAC. Traditionally, BE has been diagnosed with an invasive endoscopy of the upper gastrointestinal tract. The Cytosponge is a minimally invasive alternative to endoscopy consisting of a soluble vegetarian capsule containing a compacted sponge tethered to a thread (*5*) (**fig. 1a**). The patient swallows the capsule, and once in the proximal stomach, the capsule dissolves and the spherical sponge expands. The sponge is withdrawn using the thread and collects several million cells from the upper stomach and esophagus along its passage. Sections of tissue retrieved by the sponge are stained with H&E and trefoil factor 3 (TFF3) (**fig. 1a**), the diagnostic biomarker for intestinal metaplasia (IM) in Cytosponge-TFF3 (*6*).

**Figure 1:**
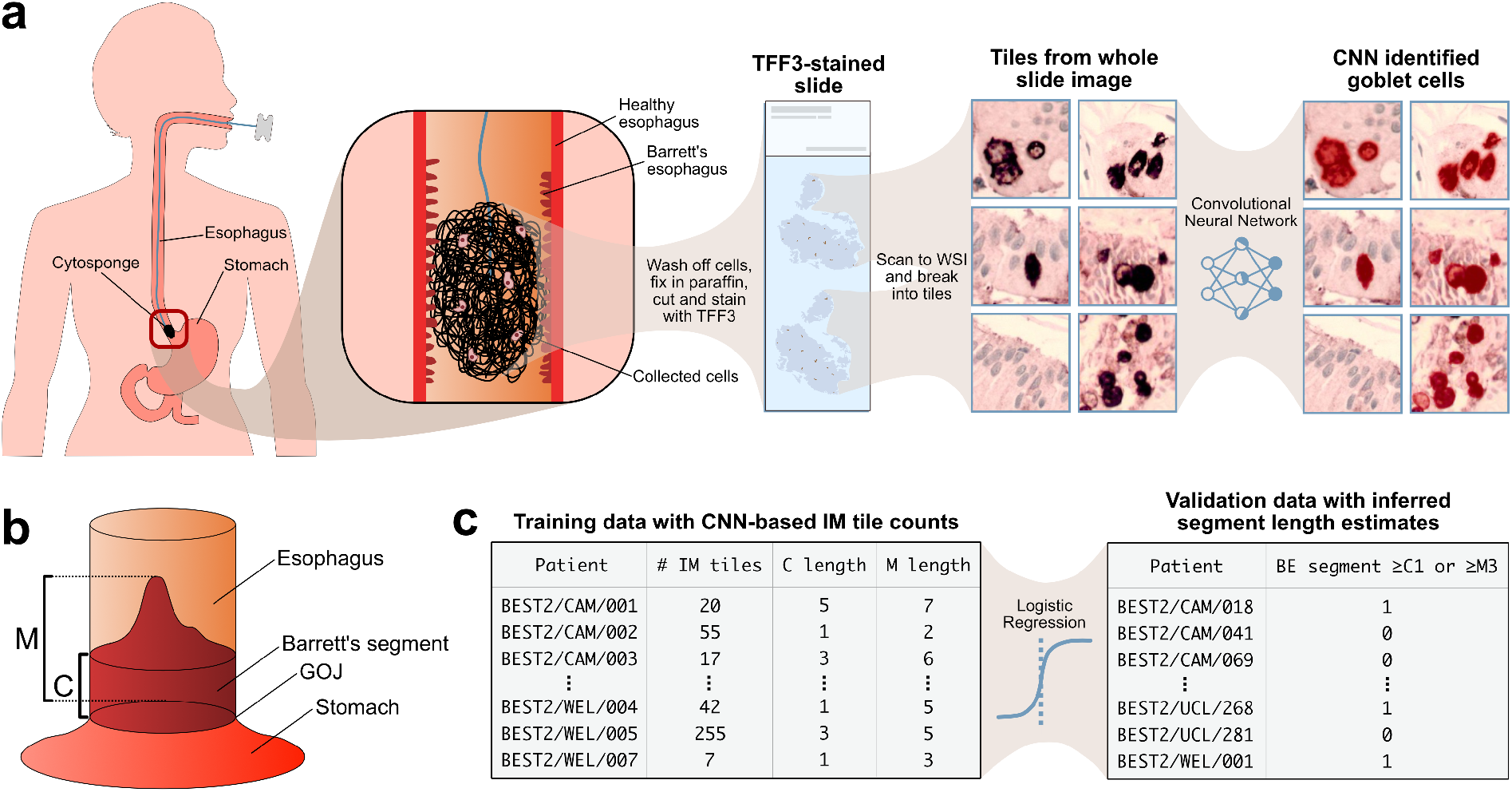
Overview of Cytosponge-TFF3 preparation, Prague classification, and computational pipeline. **(a)** The Cytosponge collects cells from the esophagus including Barrett’s cells if they are present. After the patient swallows the capsule attached to a thread, the capsule dissolves in the stomach, where the sponge spherical expands. Pulling the sponge by the thread collects cells from the upper stomach and esophagus. The cells are washed off the sponge, processed into a pellet, embedded in paraffin, cut, and stained with TFF3, and put on a slide. Slides are then scanned into whole-slide images (WSIs), which can then be broken up into thousands of smaller images (“tiles”). These tiles are labeled by a pathologist for the presence of goblet cells indicating intestinal metaplasia and a convolutional neural network is trained to perform this classification task from these labels. **(b)** A diagram showing how the C and M lengths of the Prague classification criteria for segment length measurement are found. C (circumferential) denotes distance from the proximal margin of the gastric cardial folds to the proximal margin of the circumferential BE segment, and M (maximal) describes the distance to the most proximal extent of the segment. GOJ = gastroesophageal junction. **(c)** The number of tiles classified as showing intestinal metaplasia (TFF3-positive) for slides in the training set are used to train a logistic regression model to predict the Barrett’s segment length. Length measurements are in centimeters.

Reviewing stained Cytosponge histology can be time consuming and our recent work has shown that a deep learning triage approach on a digitized whole-slide image (WSI) of the TFF3 slide (**fig. 1a**) performs the tasks of pathologists with high accuracy and can reduce pathologist workload by 57% (*7*). This was a substantial step forward to making the Cytosponge-TFF3 test widely applicable, but a major limitation remained: although BE is a precursor to cancer, not all BE cases carry the same risk (*8, 9*). Risk of cancer progression in BE can be estimated by two clinical predictors: the first one is the presence of intestinal metaplasia (IM) defined by mucin-containing cells called goblet cells, which our previous work was able to identify automatically from Cytosponge slides (*7*). The second one is the length of the BE segment, measured by the Prague classification as the circumferential (C) and maximal (M) distance the Barrett’s segment reaches from the gastroesophageal junction (*10, 11*) (**fig. 1b**). The presence of IM and the length of the Barrett’s segment dictates whether regular endoscopic surveillance or monitoring is required (*11, 12*). However, neither routine pathology review nor our novel automated predictor measure Barrett’s segment length (which is usually only attainable via endoscopy) and in all cases endoscopy follow-up is required if IM (TFF3 positivity) is detected in the Cytosponge sample.

Of all patients with a TFF3-positive Cytosponge test, 59% had a diagnosis of BE containing IM confirmed across a range of lengths (M length *<*1 to 15 cm) and in patients with no visible Barrett’s (M0), 33% had IM of the cardia identified from a single cardia biopsy (*13*). Currently, 100% of TFF3-positive patients are sent for endoscopy. Better identification of the 41% of TFF3-positive patients who do not end up having BE upon endoscopy could deprioritize a substantial fraction of TFF3-positive patients from invasive endoscopy and the large amount of resources required to perform them.

To address this challenge, we asked whether it would be possible to use machine learning on Cytosponge-TFF3 to quantify the extent of IM, and correlate that quantity with Prague length measurements (**fig. 1c**). This would provide a composite biomarker of both extent of IM and a segment length estimate from the Cytosponge-TFF3 test alone. This biomarker could have a substantial impact on patient management: patients at higher cancer risk could be prioritized for endoscopy, and those patients likely to have focal cardia IM or a very short segment could be discharged to be followed up with further Cytosponge tests without requiring an endoscopy confirmation.

To develop a composite biomarker, we took the raw output of the deep learning-based Cytosponge-TFF3 IM-prediction model (*7*) (the count of the number of tiles considered TFF3/IM-positive by the model) for whole slide images of 529 Cytosponge patients in the BEST2 trial (*14*). We correlated these counts with the Prague C and M lengths from the subsequent endoscopy. Consistent with our expectation, we found that the quantity of IM from the Cytosponge-TFF3 assay as assessed by deep learning showed a highly significant correlation with segment length. For Prague C and M lengths for the 529 TFF3-positive BEST2 patients, there was a Spearman’s rank correlation coefficient of 0.73 for C length and 0.77 for M length. A positive relationship is also visible when we plot the Prague C and M lengths against log of the TFF3-positive tile counts for the 529 patients (**fig. 2e–f**).

**Figure 2:**
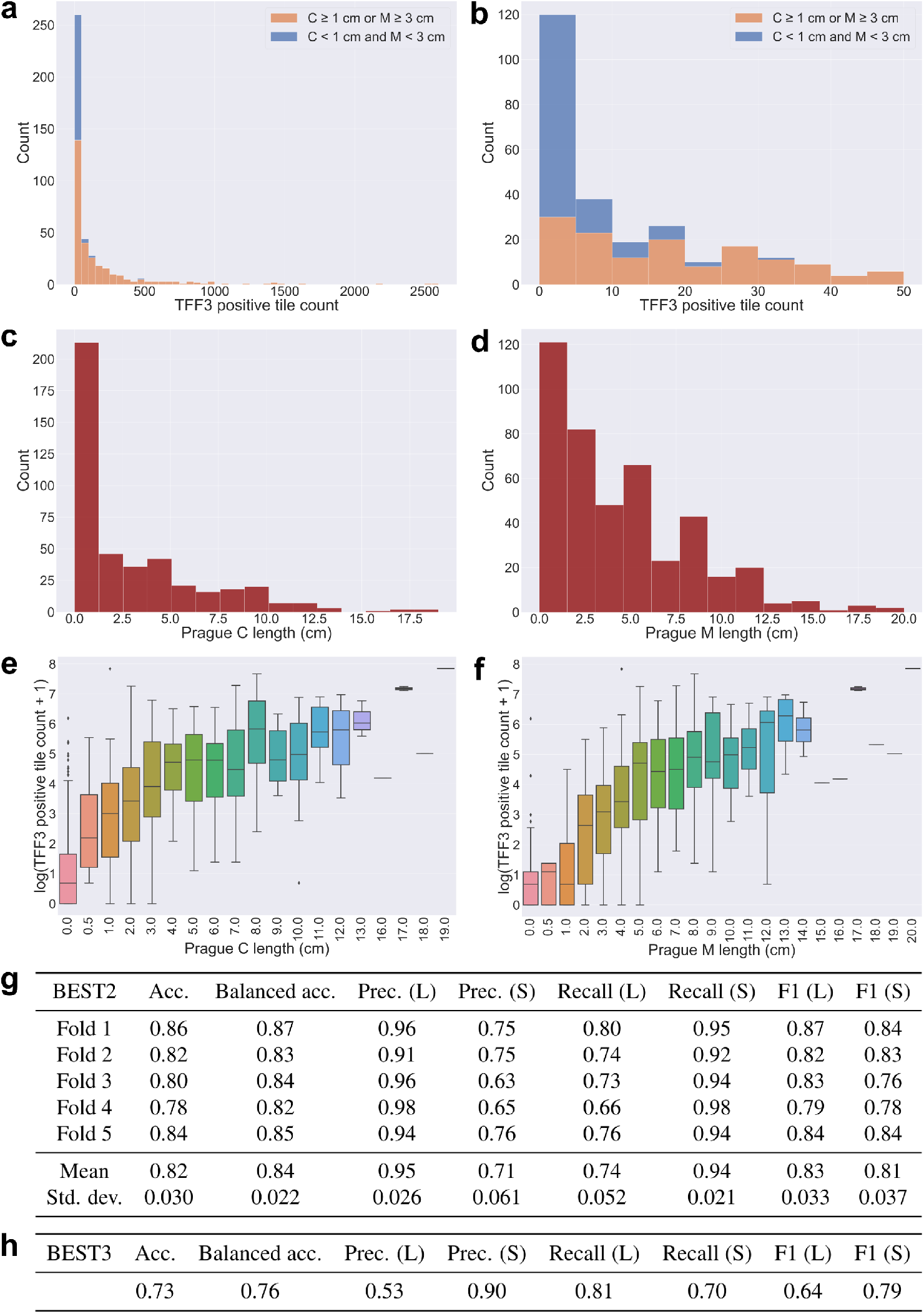
Data and logistic regression results. **(a)** A stacked-bar histogram of the machine-learning derived TFF3-positive tile counts of BEST2 patients who underwent the Cytosponge-TFF3 test. Patients with 0 TFF3-positive tiles excluded from plot. **(b)** A stacked-bar histogram like **(a)** except only showing a TFF3-positive tile count range of 0–50. Patients with 0 TFF3-positive tiles excluded from plot. **(c)** A histogram of the ground truth Prague C lengths of BEST2 patients. Patients with 0 TFF3-positive tiles excluded from plot. **(d)** A histogram of the ground truth Prague M lengths of 529 BEST2 patients. Patients with 0 TFF3-positive tiles excluded from plot. **(e)** A box-and-whiskers plot showing the log of TFF3-positive tile count vs. Prague C length for 529 BEST2 patients. **(f)** A box-and-whiskers plot showing the log of TFF3-positive tile count vs. Prague M length for 529 BEST2 patients. **(g)** Accuracy, class-balanced accuracy, precision, recall, and F1 r8esults of 5-fold cross-validation on 529 BEST2 patients applied to a logistic regression model for predicting whether or not the patient’s Barrett’s segment had C length greater than or equal to 1 cm or M length greater than or equal to 3 cm. L (“long segment”) stands for the C≥1 or M≥3 class; S (“short segment”) stands for the C*<*1 and M*<*3 class. **(h)** Results of training a model like that of **(g)**, except training on all 529 BEST2 patients without cross-validation and then inferring on a 162-patient test set of BEST3 patients. BEST3 results shown.

As a control, we took the raw output of the Cytosponge-H&E model from (*7*) for identifying tiles containing gastric tissue but no TFF3-positive tiles. As the presence of gastric tissue in a Cytosponge sample merely indicates that the device traversed through the lower esophageal sphincter into the stomach, it is expected that there should be little to no correlation between these gastric tile count and the Prague C and M lengths. Indeed, for the same 529 BEST2 patients, Spearman’s rank correlation coefficients between these tile counts and the C length was *−*0.024, and for M length was *−*0.046, confirming the null hypothesis for the control setting in comparison to IM tile count.

We next asked if the significant correlation between automatically identified IM quantity and the Barrett’s segment length could be leveraged in an optimized prediction model. We compared two strategies: one was to predict C and M lengths as continuous values (regression-based approach) and the other one was to bin C and M lengths according to clinically relevant thresholds (classification-based approach) where particular importance is placed on Barrett’s segments with C lengths greater than or equal to 1 cm, and M lengths greater than or equal to 3 cm in accordance with current Barrett’s diagnostic literature (*11, 12, 15, 16*). In both approaches the IM tile counts derived from machine learning applied to the Cytosponge-TFF3 test was the sole independent variable.

In the regression-based approach, we found that a zero-inflated Poisson (ZIP) regression model (*17*) yielded the best fit for the data (see **Methods**). We performed 5-fold cross validation on our 529 BEST2 patient cohort, all of whom underwent a Cytosponge followed by an oral endoscopy. The ZIP models achieved a mean fold *R*^2^ error of 0.40 for predicting C length and 0.40 for predicting M length (by comparison, Poisson regression without accounting for zero inflation achieves mean fold *R*^2^ errors of 0.22 for C length and 0.23 for M length). These results were not promising for clinical application.

In the classification-based approach, which takes advantage of established clinical thresholds, we trained a logistic regression model to classify whether or not a patient’s Barrett’s segment had a C length at or above 1 cm or an M length at or above 3 cm (C≥1 or M≥3 vs. C*<*1 and M*<*3). With 5-fold cross validation, the mean class-balanced validation accuracy of the logistic regression was 0.84 (0.82 unbalanced accuracy), the mean validation precision of the diagnostically positive class was 0.95, the recall 0.74, and the F1 score 0.83 (**fig. 2g**).

Next we applied this model for categorizing the segment length to the 162 patients from the BEST3 study (*13*) as an independent test set. It is important to note the differences between the clinical trials BEST2 and BEST3. BEST3 was a screening randomized controlled screening trial, in which patients on anti-reflux medication who had not been endoscoped in the past 5 years were invited for a Cytosponge test. Patients with a TFF3-positive result were invited for an endoscopy to confirm the presence of endoscopically visible columnar epithelium with IM on biopsy. It therefore includes patients with cardia IM and short Barrett’s segments who would not fulfill the guidelines for surveillance monitoring (*13*). By contrast, BEST2 was a case-control cohort study in secondary care which recruited cases with known Barrett’s esophagus undergoing surveillance (*14*). This cohort is therefore biased towards patients with longer segments requiring monitoring. Indeed, 315 of 529 BEST2 patients (59.5%) had segments with C≥1 cm or M≥3 cm, compared to just 48 of 162 BEST3 patients (29.6%). Hence, BEST2 is ideal for training the model and BEST3 for application in a screening setting in which the clinician needs to evaluate the risk to benefit ratio of performing an endoscopy to confirm Barrett’s esophagus.

When applied to patients in BEST3 with a TFF3-positive Cytosponge result the model had a precision of 0.90 for the short-segment class (80/89). The model incorrectly predicts 34 patients to have a long-segment disease however the higher number of TFF3-positive tiles may reflect more extensive intestinal metaplasia including IM of the stomach—also a pre-malignant condition (*18*). Crucially, endoscoping only the 73 patients the model predicted to have long segments (39 true positives and 34 false positives) would allow for 55% of test set patients (89/162) to avoid endoscopy while only missing 5.6% of patients with long-segment disease (9 false negatives of 162). In view of this high precision for predicting short-segment disease, and the low cancer risk associated with focal cardia IM and short-segment Barrett’s a repeat Cytosponge after an interval (e.g., three years) would seem a good alternative to endoscopy.

Our results show that not only is there a correlation between intestinal metaplasia quantification from the Cytosponge-TFF3 test and Barrett’s segment length, but that a clinically relevant segment length cutoff is predictable with high accuracy from this quantification. This establishes IM quantification as a relevant new biomarker for consideration in Barrett’s patients, as opposed to the binary classification currently used by pathologists or by the machine learning model reported previously (*7*).

In standard pathology reporting from glass slides it has not been feasible to quantify the degree of IM previously. However, since our predictions are gleaned from an entirely automated machine learning pipeline, this suggests that the incorporation of these new biomarkers into existing Barrett’s diagnosis and monitoring pathways should be readily achievable.

There is a big push towards more systematic screening for Barrett’s esophagus to improve outcomes from esophageal cancer (*8, 12–14*). In doing so, every effort must be made to maximize the benefits and reduce the harms of screening. Hence, clinical investigations that ensue from a positive screening result should spare patients at very low risk for cancer and be affordable for the health care system. If Cytosponge was adopted for screening on a population scale, deep-learning based IM quantification from Cytosponge-TFF3 and the accompanying Barrett’s segment prediction could help focus endoscopy on those who need it most. A similar approach could also be applied to Barrett’s surveillance procedures using Cytosponge or endoscopic biopsies to quantify the degree of IM as part of a cancer risk score. Overall, these results are progress towards stratifying patients at increased cancer risk for expedient endoscopy while saving lower-risk patients from more invasive procedures as well as the resources for performing them.

## Methods

The deep learning model to elucidate the number TFF3-positive tiles comes from a deep learning pipeline described in full in (*7*), and making use of PathML to perform the tiling task (*19*). Cytosponge samples with an H&E slide showing inadequate amounts of gastric tissue (as determined automatically in (*7*)) were excluded. The logistic regression model for predicting segment length from IM tile count was implemented with the `glm()` function in the R programming language, and the Spearman’s correlation was performed with R’s `cor()` function. Zero-inflated Poisson regression was performed with R’s `pscl` library (*20*). Plots were generated with Python’s `matplotlib` library.

Documented code with instructions for running the analyses shown in this paper, including generating diagnostic plots for the regression model, can be found at the following public repository: https://github.com/markowetzlab/barretts-segment-length-predictor.

## Data Availability

The data and fully documented code to perform all of the analyses of this paper can be found at this public repository in the form of two illustrative Jupyter notebooks: https://github.com/markowetzlab/barretts-segment-length-predictor

## Acknowledgments

We would like to thank William Orchard for his assistance on the regression model, as well as Marcel Gehrung for producing the model from which TFF3-positive tile counts were derived. We would also like to thank the pathologists of Addenbrooke’s Hospital for their previous scoring of Cytosponge-TFF3 and Cytosponge-H&E slides as part of the BEST clinical trials. This research was supported by Cancer Research UK (FM: C14303/A17197), a Medical Research Council programme grant (RCF: RG84369) and Cambridge University Hospitals NHS Foundation Trust. BEST2 was funded by Cancer Research UK (12088 and 16893). We would also like to thank the NIHR Cambridge Biomedical Research Centre (BRC-1215-20014) and the Experimental Cancer Medicine Centre for their support. AGB acknowledges support from a Gates Cambridge Scholarship from the Bill & Melinda Gates Foundation. FM is a Royal Society Wolfson Research Merit Award holder.

## Author contributions

RCF conceived the study. AGB implemented the analyses. All work was performed under the supervision of FM and RCF The final manuscript was read and approved by all authors.

## Declaration of interests

R.C.F is named on patents related to Cytosponge-TFF3 that have been licensed by the Medical Research Council to Covidien (now Medtronic). RCF is a founder and shareholder of Cyted Ltd. FM is a founder, director and shareholder of Tailor Bio.

## Notes

### Clinical Trial

ISRCTN12730505, ISRCTN68382401

### Clinical Protocols

https://github.com/markowetzlab/barretts-segment-length-predictor

### Author Declarations

BEST2: Ethics approval was obtained from the East of England - Cambridge Central Research Ethics Committee (number 10/H0308/71) and registered in the UK Clinical Research Network Study Portfolio (9461). BEST3: Ethics approval was obtained from the East of England - Cambridge Central Research Ethics Committee (number 16/EE/0546).

## References

1. Bray, F. et al. Global cancer statistics 2018: Globocan estimates of incidence and mortality worldwide for 36 cancers in 185 countries. CA: a cancer journal for clinicians 68, 394–424 (2018).

2. Pohl, H., Sirovich, B. & Welch, H. G. Esophageal adenocarcinoma incidence: are we reaching the peak? Cancer Epidemiology and Prevention Biomarkers 19, 1468–1470 (2010).

3. Smyth, E. C. et al. Oesophageal cancer. Nature reviews Disease primers 3, 17048 (2017).

4. Peters, Y. et al. Barrett oesophagus. Nature Reviews Disease Primers 5 (2019). URL https://doi.org/10.1038/s41572-019-0086-z.

5. Ross-Innes, C. S. et al. Evaluation of a minimally invasive cell sampling device coupled with assessment of trefoil factor 3 expression for diagnosing barrett’s esophagus: a multi-center case– control study. PLoS medicine 12, e1001780 (2015).

6. Paterson, A. L., Gehrung, M., Fitzgerald, R. C. & O’Donovan, M. Role of tff3 as an adjunct in the diagnosis of barrett’s esophagus using a minimally invasive esophageal sampling device—the cytosponge. Diagnostic Cytopathology (2019). URL https://onlinelibrary.wiley.com/doi/abs/10.1002/dc.24354. https://onlinelibrary.wiley.com/doi/pdf/10.1002/dc.24354.

7. Gehrung, M. et al. Triage-driven diagnosis of barrett’s esophagus for early detection of esophageal adenocarcinoma using deep learning. Nature Medicine 27, 833–841 (2021).

8. Sharma, P. et al. Dysplasia and cancer in a large multicenter cohort of patients with barrett’s esophagus. Clinical Gastroenterology and Hepatology 4, 566–572 (2006).

9. Bhat, S. et al. Risk of malignant progression in barrett’s esophagus patients: Results from a large population-based study. Journal of the National Cancer Institute 103, 1049–1057 (2011).

10. Sharma, P. et al. The development and validation of an endoscopic grading system for barrett’s esophagus: the prague c & m criteria. Gastroenterology 131, 1392–1399 (2006).

11. Shaheen, N. J., Falk, G. W., Iyer, P. G. & Gerson, L. B. Acg clinical guideline: Diagnosis and management of barrett’s esophagus. American Journal of Gastroenterology 111, 30–50 (2016).

12. Fitzgerald, R. C. et al. British society of gastroenterology guidelines on the diagnosis and management of barrett’s oesophagus. Gut 63, 7–42 (2018).

13. Fitzgerald, R. C. et al. Cytosponge-trefoil factor 3 versus usual care to identify barrett’s oesophagus in a primary care setting: a multicentre, pragmatic, randomised controlled trial. The Lancet 396, 333–344 (2020).

14. Ross-Innes, C. et al. Risk stratification of barrett’s oesophagus using a non-endoscopic sampling method coupled with a biomarker panel: a cohort study. Lancet Gastroenterol Hepatol. 2, 23–31 (2017).

15. Parasa, S. et al. Development and validation of a model to determine risk of progression of barrett’s esophagus to neoplasia. Gastroenterology 154, 1282–1289.e2 (2018).

16. Qumseya, B. et al. Asge guideline on screening and surveillance of barrett’s esophagus. Gastrointestinal Endoscopy 90, 335–359.e2 (2019).

17. Lambert, D. Zero-inflated poisson regression, with an application to defects in manufacturing. Technometrics 34, 1–14 (1992).

18. Hadjinicolaou, A. et al. Cytosponge-tff3 testing can detect precancerous mucosal changes of the stomach. Clinical Gastroenterology and Hepatology In press (2021).

19. Berman, A. G., Orchard, W. R., Gehrung, M. & Markowetz, F. Pathml: A unified framework for whole-slide image analysis with deep learning. medRxiv (2021). URL https://www.medrxiv.org/content/early/2021/07/13/2021.07.07.21260138. https://www.medrxiv.org/content/early/2021/07/13/2021.07.07.21260138.full.pdf.

20. Zeileis, A., Kleiber, C. & Jackman, S. Regression models for count data in R. Journal of Statistical Software 27 (2008). URL http://www.jstatsoft.org/v27/i08/.

